# Psychiatric morbidity and protracted symptoms in recovered COVID-19 patients

**DOI:** 10.1101/2020.10.07.20208249

**Authors:** Burç Çağrı Poyraz, Cana Aksoy Poyraz, Yeşim Olğun, Özge Gürel, Sena Alkan, Yusuf Emre Özdemir, İlker İnanç Balkan, Rıdvan Karaali

## Abstract

We investigated the psychiatric symptomatology and the protracted symptoms in recently recovered COVID-19 patients. This cross-sectional study assessed 284 patients recruited from a tertiary hospital. Patients completed a web-based survey on socio-demographic data, past medical/psychiatric history, and additional information relevant to the outbreak conditions. The psychiatric status was assessed using the Impact of Events Scale-Revised (IES-R), Hospital Anxiety and Depression Scale (HADS), Pittsburgh Sleep Quality Index (PSQI), and MINI suicidality scale. Patients completed a checklist for the acute symptom burden and protracted symptoms that were experienced after the acute infection. After a mean of 50 days following the diagnosis of COVID-19, 98 patients (34.5%) reported clinically significant PTSD, anxiety, and/or depression, with PTSD being the most common condition reported (25.4%). One hundred and eighteen patients (44.3%) reported one or more protracted symptom(s), with fatigue, muscle aches, alteration of smell/taste, headache and difficulty in concentration, being the most common symptoms reported. Predictors of PTSD symptom severity were the female gender, past traumatic events, protracted symptoms, perceived stigmatization, and a negative view on the seriousness of the COVID-19 pandemic. Binary logistic regression analysis showed that PTSD symptom severity was the sole independent predictor of the presence of protracted symptoms. Our results suggest that COVID-19 patients may be prone to substantial psychological distress in the first months after the infection. The protracted symptoms were also frequent in this period, and these were related to the posttraumatic psychiatric morbidity. Both the psychiatric morbidity and the protracted symptoms were independent of the initial infection severity. Further research on the neurobiological commonalities between the protracted symptoms and PTSD in COVID-19 patients is warranted.

## 1. Introduction

The novel coronavirus disease (COVID-19) arose in Wuhan, China in December 2019 and spread worldwide rapidly, with the declaration of a global pandemic by the World Health Organization (WHO) as of March 2020. As the COVID-19 pandemic progresses worldwide, its psychological impact is increasingly being recognized among the vulnerable groups, including the health care workers, individuals on quarantine, patients with chronic medical diseases and psychiatric disorders, as well as the public in general.

Only a few studies have investigated the psychiatric sequelae related to the COVID-19 infection. While delirium, insomnia, and symptoms of depression, anxiety, and posttraumatic stress disorder (PTSD) have been reported as common features in the acute (and immediate convalescent) period, data are sparse on the psychiatric status in the post-illness phase (Rogers et al. 2020). To our knowledge, only three studies investigated the psychiatric symptomatology in the recovered COVID-19 patients, and these reported high rates of insomnia, and symptoms of PTSD, depression, and anxiety, about one month after the infection (Liu et al. 2020; Mazza et al. 2020; Tomasoni et al. 2020). These preliminary findings suggest substantial psychiatric morbidity following the COVID-19 infection, comparable to what was reported after the previous coronavirus epidemics (Severe Acute Respiratory Syndrome (SARS) and Middle East Respiratory Syndrome (MERS) (Wu et al. 2020; Mak et al. 2009; Lee et al. 2007).

A recent concern for the patients who had recovered from the COVID-19 infection is the protracted symptoms, such as chronic fatigue, diffuse myalgia, dyspnea, headache, and concentration difficulties which may result in significant disability (Carfi et al. 2020; Townsend et al. 2020). Interestingly, although follow-up time was longer, a similar course of symptoms has been implicated in the previous coronavirus outbreaks (SARS and MERS) (Lam et al. 2009; Lee et al. 2019; Moldofsky and Patcai 2011). Although the clinical similarities of this phenomenon with the postinfectious chronic fatigue syndrome have been argued, the etiopathogenesis is far from clear, and the relation of these protracted symptoms with the psychiatric status has not been investigated.

The present study aimed to examine the extent of the psychiatric symptomatology (symptoms of PTSD, anxiety and depression, sleep impairment, and suicidality) in the recovered COVID-19 patients. We also evaluated the potential predictors of the significant PTSD symptoms, based on a theoretical susceptibility model: the individual pre-trauma and peri-trauma factors (Sareen, 2014) Lastly, we aimed at exploring the prevalence of protracted symptoms and interrelations of these symptoms with the current psychiatric status.

## 2. Methods

### 2.1. Patients and procedure

We performed a cross-sectional survey study investigating the psychological well-being of patients with ‘probable’ and ‘confirmed’ diagnosis of the COVID-19 infection (ECDPC criteria, 2020), after completion of their medical care. The study population included the adult patients who had received care at the tertiary hospital of Cerrahpaşa Medical Faculty, Istanbul, between March 15 and May 15, 2020. A total of 1200 patients meeting the WHO criteria for discontinuation of quarantine (no fever in three consecutive days and 14 days after significant clinical improvement) were identified from the hospital records, and were contacted via WhatsApp^®^ and short message service (SMS) messages (WHO Criteria, 2020). These were invited to participate in the online survey designed via the Survey Monkey^®^ online survey portal. We also enrolled volunteering post-acute COVID-19 outpatients followed by the infectious disease department of the above hospital. An identical paper survey was administered to the latter group. The study was performed between June 1 and July 1, 2020. The main areas of interest were the PTSD symptoms, symptoms of anxiety and depression, sleep impairment, suicidality, and the protracted symptoms. We also investigated the potential predictors of the PTSD symptoms in the sample based on a theoretical susceptibility model: the pre-trauma (i.e. demographic characteristics, psychiatric and medical history, past traumatic experiences) and peri-trauma factors (i.e. severity of the COVID-19 infection, hospitalization status, protracted symptoms, COVID-19 infection in the social vicinity, social support, stigmatization, and job loss during the outbreak).

In response to WhatsApp^®^ and SMS invitations, 239 responses were received (response rate = 20%). Additionally, 79 paper surveys were completed by the volunteering outpatients.

#### None

Eventually, 284 surveys with a complete IES-R were included in the analysis. On average, respondents spent 22 minutes on the online survey.

An informed consent form was presented on the first page of the (electronic or written) survey citing the purposes and the voluntary nature of the survey, and that all information provided by the participants would be kept confidential, and they could withdraw from the survey at any time. The procedures of this study complied with the provisions of the Declaration of Helsinki concerning research on Human participants. The Ethics Committee of Cerrahpaşa Medical Faculty approved this study.

### 2.2. Data collection, demographic, social and clinical parameters

#### 2.2.1 Questionnaire on the demographic status and the general effects of the outbreak

The survey contained three parts and 128 questions. The first part included a self-report questionnaire to collect sociodemographic variables of interest (age, gender, educational, marital and employment status, monthly family income, and household size), medical status (history of chronic diseases and psychiatric disorders), and additional information relevant to the outbreak and lockdown conditions (having a child below 18 years of age, the presence of an individual in the household above 65 years of age or with significant medical risks related to the COVID-19, family history of COVID-19, history of COVID-19 in patient’s relatives, friends and acquaintances, change in patient’s employment status during the outbreak, patient’s sources of information during the outbreak (media type), duration of daily TV and social media exposure, and personal view on the seriousness of the COVID-19 outbreak: ‘a very serious threat’, ‘a serious threat’, ‘a small threat’ or ‘not a real threat’).

#### 2.2.2 Questionnaire on patients’ history of the COVID-19 infection

The second part of the survey included questions on the patient’s past COVID-19 infection. We questioned the date the initial symptoms of the infection appeared, a range of acute infectious symptoms experienced by the patient (i.e. fever, cough, malaise, dyspnea, sore throat, rhinorrhea, lightheadedness, headache, nausea, diarrhea, abdominal pain, muscle aches, alteration of the senses of smell or taste, numbness and tingling sensations on the skin, difficulty in concentration, and daytime sleepiness), the duration of these symptoms, whether the patient was hospitalized or followed on an outpatient basis, and a personal view on the general severity of the infectious symptoms experienced 0 (no symptoms), 1 (very mild symptoms), 2 (mild symptoms), 3 (moderate symptoms), 4 (severe symptoms), or 5 (very severe symptoms). We also questioned on a checklist whether the potential symptoms of interest persisted after the acute infectious symptoms subsided. These symptoms included the alteration of smell and taste, headache, fatigue, daytime sleepiness, muscle aches, lightheadedness, difficulty in concentration, and numbness and tingling sensations on the skin.

#### 2.2.3. Review of patients’ medical files

Patients’ medical files were reviewed for the relevant information regarding the COVID-19 clinical severity scoring (asymptomatic, mild, moderate, severe and critical disease; WHO criteria, 2020), lowest oxygen saturation levels on pulse oximetry, whether supplemental oxygen was required during treatment, and the experimental agents administered.

### 2.3. Psychometric assessment

#### 2.3.1 Impact of Event Scale-Revised (IES-R)

Symptoms of PTSD related to the outbreak were assessed using the Impact of Event Scale-Revised (IES-R) (Weiss and Marmar, 1997). This tool is a 22-item self-report questionnaire that evaluates symptoms of intrusion, avoidance, and hyperarousal, and presents a total score for the subjective stress related to a traumatic event. IES-R was frequently used after a variety of traumatic settings (Morina et al., 2013), and after major public health crises (Lee et al., 2018; Varshney et al., 2020; Wang et al., 2020). We made slight modifications to the Turkish adaptation of the IES-R (Corapcioglu et al., 2006) (replacing the word ‘event’ with ‘outbreak’, where appropriate) in order to account for the nature of the event investigated.

#### 2.3.2. Hospital Anxiety and Depression Scale (HADS)

Anxious and depressive symptoms were assessed using the Hospital Anxiety and Depression Scale (HADS) (Zigmond and Snaith, 1983), a 14-item self-report questionnaire. HADS can assess symptom severity and caseness of anxiety and depressive disorders, and it consists of anxiety and depression subscales. HADS was validated in a variety of populations, including the general medical and community settings (Bjelland et al., 2002; Bocéréan and Dupret, 2014; Djukanovic et al., 2017). HADS was validated previously for the Turkish population (Aydemir et al., 1997).

#### 2.3.3. Life Events Checklist (LEC)

The Life Events Checklist (LEC) was used to screen for the potentially traumatic events in the subject’s lifetime. (Gray et al., 2004) LEC is a 17-item, self-report measure that assesses exposure to 16 events known to potentially lead to PTSD, and one other extraordinarily stressful event not captured in the list. A past traumatic experiences score was calculated for each subject based on the number of following items checked by the subject (i.e. “happened to me”, “witnessed it”, “learned about it” and “part of my job”), with a possible range of 0 to 68 points.

#### 2.3.4. Pittsburgh Sleep Quality Index (PSQI)

Sleep impairment in the prior month was assessed by selected items of the Turkish version of the Pittsburgh Sleep Quality Index (PSQI), a self-report questionnaire that assesses a broad range of sleep dimensions.(Buysse et al., 1989) In this study, patients’ sleep latency (question 2 and 5a), sleep duration (question 4), sleep disturbances (questions 5b-5j), and subjective sleep quality (question 9) were scored according to PSQI, each from 0 to 3 with higher scores indicating worse sleep quality. PSQI has been validated for the Turkish population (Ağargün et al., 1996)

#### 2.3.5. MINI suicidality scale

Suicidality was assessed following the short version of the MINI Suicidality Scale, which consists of six items scored as ‘yes’ or ‘no’: intentions of self-harm, wish for death, thoughts of suicide, suicidal plan, attempted suicide in the past month, and lifetime history of a suicide attempt. The combined suicidality score is calculated based on the weighted score of each item, with a range of 0–33 points. (Sheehan, D. V. 1998)

### 2.4. Stigmatization and social support

The patients rated how much they felt they were stigmatized and discriminated against, on a scale of 0 (never), 1 (very little), 2 (moderately), or 3 (considerably). They also rated the support from the family and friends on a scale of 0 (no support), 1 (little support), 2 (moderate support), or 3 (considerable support).

### 2.5. Statistical Analysis

We subdivided our sample into three groups according to total IES-R score: 0-23 (normal), 24-32 (mild PTSD symptoms), and 33 and above (moderate-to-severe PTSD symptoms). Univariate analyses were made for comparison of continuous (ANOVA) and categorical variables (contingency table/X^2^) in the three groups. Chosen significant variables subsequently underwent an ordinal logistic regression to evaluate the impact of independent variables on the ordinal categories of PTSD symptom severity. We also performed binary logistic regression analysis to explore the predictors of persistence of symptoms, using the significant independent variables found in the univariate analyses. All tests were two-tailed, with a significance level of p < 0.05. Statistical analysis was performed using SPSS Statistic 23.0 (IBM SPSS Statistics, New York, United States).

## 3. Results

### 3.1. PTSD, depression, anxiety, sleep, and the epidemiological data

Epidemiological characteristics and results of the analyses of these data across comparison groups (normal, mild, and moderate-to-severe PTSD symptoms) are summarized in Table 1. Subjects’ (N=284) mean age was 39.7 (SD=12.7), and females constituted 49.8% of the sample. The majority of subjects were between 28-57 years of age (69.4%), married (65%), employed (68.3%), had a university or higher education (50%), had a child less than 18 years of age (65.3%), and had a household size of 3 or 4 individuals (54.3%).

**Table 1.** Epidemiological characteristics and the analyses of these data across three comparison groups (normal, mild, and moderate-to-severe PTSD symptoms)

| *Characteristics n (%); mean (SD)* | **Total** | **Normal** | **Mild PTSD symptoms** | **Moderate-to-severe PTSD symptoms** | **X<sup>2</sup>/F** | **df** | **p** |
| --- | --- | --- | --- | --- | --- | --- | --- |
| **Overall** | 284 | 160 (56.3) | 52 (18.3) | 72 (25.4) |  |  |  |
| **Age** |  |  |  |  | 5.54 | 8 | 0.69 |
| 18-27 | 64 (23.4) | 35 (22.6) | 10 (19.6) | 19 (27.9) |  |  |  |
| 28-37 | 58 (21.2) | 38 (24.5) | 11 (21.6) | 9 (13.2) |  |  |  |
| 38-47 | 68 (24.8) | 36 (23.2) | 13 (25.5) | 19 (27.9) |  |  |  |
| 48-57 | 64 (23.4) | 33 (21.3) | 14 (27.5) | 17 (25) |  |  |  |
| 57+ | 20 (7.3) | 13 (8.4) | 3 (5.9) | 4 (5.9) |  |  |  |
| **Gender** |  |  |  |  | 17.13 | 2 | <0.001 |
| Male | 140 (50.2) | 96 (60.8) | 21 (41.2) | 23 (32.9) |  |  |  |
| Female | 139 (49.8) | 62 (39.2) | 30 (58.8) | 47 (67.1) |  |  |  |
| **Educational status** |  |  |  |  | 5.92 | 6 | 0.43 |
| Primary | 57 (20.5) | 29 (18.5) | 12 (23.5) | 16 (22.9) |  |  |  |
| Lower secondary | 18 (6.5) | 10 (6.4) | 5 (9.8) | 3 (4.3) |  |  |  |
| Upper secondary | 64 (23) | 40 (25.5) | 6 (11.8) | 18 (25.7) |  |  |  |
| University or higher | 139 (50) | 78 (49.7) | 28 (54.9) | 33 (47.1) |  |  |  |
| **Occupation** |  |  |  |  | 13.81 | 6 | 0.03 |
| Employed |  |  |  |  |  |  |  |
| Health sector | 76 (27.6) | 42 (27.1) | 19 (36.5) | 15 (22.1) |  |  |  |
| Other | 112 (40.7) | 72 (46.5) | 19 (36.5) | 21 (30.9) |  |  |  |
| Unemployed | 69 (25.1) | 31 (20) | 13 (25) | 25 (36.8) |  |  |  |
| Student | 18 (6.5) | 10 (6.5) | 1 (1.9) | 7 (10.3) |  |  |  |
| **Marital status** |  |  |  |  | 1.80 | 2 | 0.40 |
| Married | 184 (65) | 109 (68.1) | 33 (63.5) | 42 (59.2) |  |  |  |
| Not married | 99 (35) | 51 (31.9) | 19 (36.5) | 29 (40.8) |  |  |  |
| **Family income (TL)** | 8.029 (9.838) | 8.819 (10.987) | 8.486 (10.790) | 6.040 (5.436) | 1.85 | 2 | 0.15 |
| **Household size** |  |  |  |  | 4.43 | 4 | 0.35 |
| Less than 3 individuals | 56 (20) | 31 (19.5) | 11 (21.2) | 14 (20.3) |  |  |  |
| 3-4 individuals | 152 (54.3) | 94 (59.1) | 24 (46.2) | 34 (49.3) |  |  |  |
| >4 individuals | 72 (25.7) | 34 (21.4) | 17 (32.7) | 21 (30.4) |  |  |  |
| **Has a child less than 18 years** | 181 (65.3) | 101 (65.2) | 36 (69.2) | 44 (62.9) | 0.54 | 2 | 0.76 |
| **Individual above 65 years in household** | 78 (27.9) | 46 (29.3) | 9 (17.3) | 23 (32.4) | 3.76 | 2 | 0.15 |

The mean sample score for the IES-R was 22.2 (SD=14.8; range=0-72). In our sample, 72 subjects (25.4%) reported moderate-to-severe PTSD symptoms (or clinical ‘caseness’; IES-R score equal to or above 33), while 52 subjects (18.3%) had mild PTSD symptoms (or ‘partial’ PTSD; IES-R score equal to or above 24, Creamer et al., 2003)).

Mean sample scores for HADS anxiety and HADS depression were 6.2 (SD=4.6; range=0-21) and 6.3 (SD=4.3; range=0-21), respectively. Identification of ‘probable’ cases of anxiety and depression was based on the cutoff scores recommended by the authors of the HADS scale (Zigmond and Snaith, 1983). Fifty subjects (18.4%) were considered having ‘probable’ anxiety (HADS anxiety score above 10), and 51 subjects (18.8%) were considered having ‘probable’ depression (HADS depression score above 10).

Overall, of the 72 subjects reporting moderate-to-severe PTSD symptoms, 37 subjects (51.3%) reported comorbid anxiety, and 29 subjects (40.2%) reported comorbid depression. Of all the patients, 98 (34.5%) had either moderate-to-severe PTSD, probable anxiety, or probable depression.

One hundred and one subjects (38.8%) had a subjective sleep quality rating of a ‘fairly poor or ‘very poor sleep’ in the previous month. Fifty-four subjects (22.4%) had a sleep duration of 5-6 hours or shorter, and 46 subjects (18.8%) had a sleep latency of an hour or longer. Overall, of the subjects who reported moderate-to-severe PTSD symptoms, 45 subjects (68.2%) reported poor sleep (fairly or very poor sleep) in the previous month.

Twenty-one subjects (9.9%) had a positive response to one or more items in the MINI suicidality scale, and six of these subjects (2.8%) had a ‘moderate’ current risk of suicide, based on MINI combined score (a score of 6-9). Overall, of the subjects who reported moderate-to-severe PTSD symptoms, 13 subjects (23.2%) had a positive response to one or more items, and the current risk of suicide was ‘moderate’ for four of them (7.1%). None of the subjects in our sample had a ‘high’ current risk of suicide, based on MINI combined score (a score of 10 or above).

A chi-square test (gender X PTSD severity) showed that PTSD severity differed by gender (*X*^2^ (2, *N* = 284) = 17.27, p<0.001), and a significantly higher number of subjects reporting no PTSD symptoms were males (p<0.05, post hoc analyses with Bonferroni correction).

Another chi-square test (occupation X PTSD severity) showed that PTSD severity also differed by the occupation (*X*^2^ (2, *N* = 275) = 13.81, p=0.03), and a significantly higher number of subjects reporting moderate-to-severe PTSD symptoms were unemployed (including housewives) (p<0.05, post hoc analyses with Bonferroni correction).

Age, educational status, marital status, family income, household size, the presence of an individual above 65 years of age in the household or with significant medical risks related to COVID-19, and the status of having a child less than 18 years of age did not differ among the comparison groups.

### 3.2. Data related to the COVID-19 infection

COVID-19 infection-related data and results of the analyses of these data across comparison groups (normal, mild, and moderate-to-severe PTSD symptoms) are summarized in Table 2.

**Table 2.** COVID-19 infection-related data and results of the analyses of these data across three comparison groups (normal, mild, and moderate-to-severe PTSD symptoms)

| *Characteristics n (%);*<br>*Mean (SD)* | **Total** | **Normal** | **Mild PTSD**<br>**symptoms** | **Moderate-to-**<br>**severe PTSD**<br>**symptoms** | **X<sup>2</sup>/F** | **df** | **p** |
| --- | --- | --- | --- | --- | --- | --- | --- |
| **Overall** | 284 | 160 (56.3) | 52 (18.3) | 72 (25.4) |  |  |  |
| **COVID-19 Severity** |  |  |  |  | 0.63 | 2 | 0.53 |
| Asymptomatic | 7 (2.8) | 7 (5.1) | - | - |  |  |  |
| Mild | 125 (50.8) | 69 (50.7) | 18 (40.9) | 38 (57.6) |  |  |  |
| Moderate | 80 (32.5) | 40 (29.4) | 21 (47.7) | 19 (28.8) |  |  |  |
| Severe | 31 (12.6) | 18 (13.2) | 5 (11.4) | 8 (12.1) |  |  |  |
| Critical | 3 (1.2) | 2 (1.5) | - | 1 (1.5) |  |  |  |
| **Supplemental oxygen required during**<br>**treatment** | 52 (21.2) | 30 (22.1) | 10 (22.7) | 12 (18.5) | 0.41 | 2 | 0.81 |
| **Lowest saturation level** | 95.7 (3.9) | 95.7 (4) | 95.4 (3.4) | 95.6 (4) | 0.12 | 2 | 0.88 |
| **Chronic medical disease** | 92 (34.2) | 47 (31.1) | 18 (37.5) | 27 (38.6) | 1.46 | 2 | 0.48 |
| **Treatment setting** |  |  |  |  | 1.76 | 2 | 0.41 |
| Hospitalized | 112 (39.9) | 59 (36.9) | 24 (47.1) | 29 (41.4) |  |  |  |
| Outpatient | 169 (60.1) | 101 (63.1) | 27 (52.9) | 41 (58.6) |  |  |  |
| **Acute infectious symptom burden**<br>**(# of symptom s)** | 5.3 (3.0) | 4.7 (2.7) | 5.9 (3.1) | 6.3 (3.3) | 8.36 | 2 | <0.001 |
| **How long did your acute infectious**<br>**symptoms last? (days)** | 13.4 (9.3) | 11.2 (8.5) | 15.3 (8.8) | 16.7 (10.3) | 9.16 | 2 | <0.001 |
| **How severe were your acute infectious**<br>**symptoms?** |  |  |  |  | 20.36 | 4 | <0.001 |
| “No symptoms”, “Very Mild” or “Mild” | 93 (33.2) | 69 (43.7) | 12 (23.5) | 12 (16.9) |  |  |  |
| “Moderate” | 89 (31.8) | 46 (29.1) | 19 (37.3) | 24 (33.8) |  |  |  |
| “Severe” or “Very severe” | 98 (35) | 43 (27.2) | 20 (29.2) | 35 (49.3) |  |  |  |
| **Protracted symptoms**<br>**(# of symptom s)** | 1.5 (1.6) | 0.9 (1.3) | 1.8 (1.6) | 2.4 (1.9) | 21.32 | 2 | <0.001 |

The time between the diagnosis of COVID-19 infection and the survey response was 48.7 days (SD = 20.4; range = 14-116 days). Two hundred and twenty subjects (88%) had a PCR confirmed diagnosis, while 30 subjects (12%) were ‘probable’ patients based on the clinical and CT findings.

The COVID-19 infection severity among subjects based on the WHO criteria is depicted in Table 2. Of the subjects, 60.1% were followed on an outpatient basis, and 39.9% were hospitalized. Three subjects had been admitted to the intensive care unit (ICU). Of 245 patients who had oxygen saturation records based on the pulse oximeter, 51 (20.8%) had a saturation below 94. Similarly, 52 subjects (21.2%) required oxygen treatment during their hospitalization. The most common experimental treatment used was hydroxychloroquine (229 patients; 92.3%), followed by oseltamivir (104 patients; 41.9%) and azithromycin (75 patients; 30.2%). Sixty patients (24.5%) received favipiravir treatment, and 14 patients (5.7%) were treated with tocilizumab.

The infection severity (WHO criteria, 2020), the status of having been hospitalized, oxygen saturation levels, and the need for supplemental oxygen during hospitalization did not differ among the comparison groups based on the PTSD severity. Also, the frequency of each experimental agent used was not statistically different between comparison groups.

Ninety-two patients (34.2%) had one or more chronic medical disease(s). Among these, hypertension (10.4%), diabetes mellitus (8.6%), cardiac diseases (9.7%), pulmonary diseases (8.2%), and cancer (3%) were the most common diagnoses. A chi-square test (chronic medical disease X PTSD severity) showed that PTSD severity did not differ by the presence of chronic medical disease (*X*^2^ (2, *N* = 269) = 1.46, p=0.48).

Patients reported that they experienced a median of 5 symptoms (range=0-14) during the acute COVID-19 infection, with malaise (78%), muscle aches (61%), headache (52%), alteration of taste (54%), fever (52%), alteration of smell (48%), cough (47%), diarrhea (35%), sore throat (33%), dyspnea (32%), daytime sleepiness (30%), nausea (29%), rhinorrhea (22%), difficulty in concentration (28.3%), lightheadedness (28%), numbness and tingling sensations on the skin (12%), and abdominal pain (12%), being the common symptoms reported. Patients also reported that their acute symptoms lasted a median of 10 days (range=0-50). Ninety-three subjects (33.2%) reported that their COVID-19 infection was ‘very mild’, ‘mild’, or they did not have any symptoms. For 89 subjects (31.8%), the severity of symptoms was ‘moderate’, and for 98 patients (35%), the symptoms were ‘severe’ or ‘very severe’.

ANOVAs revealed significant differences among the comparison groups (normal, mild and moderate-to-severe PTSD symptoms) regarding the reported acute symptom burden (F (2, 270) = 8.36; p < 0.001), and the reported length of acute symptoms (F (2, 245) = 9.16; p < 0.001). Post hoc Tukey HSD tests indicated that the mean acute symptom burden for the moderate-to-severe and mild PTSD groups were significantly higher than no PTSD group. Also, post hoc Tukey HSD tests indicated that the mean length of acute symptoms for the moderate-to-severe and mild PTSD groups were significantly higher than no PTSD group.

A chi-square test (reported COVID-19 infection severity X PTSD severity) showed that the reported severity of the COVID-19 infection differed by the PTSD severity (*X*^2^ (4, *N* = 280) = 20.39, p<0.001), and a report of asymptomatic infection, and ‘mild’ or ‘very mild’ symptoms was significantly more common in patients reporting no PTSD symptoms.

One hundred and eighteen patients (44.3%) reported one or more potential symptom(s) that persisted after the acute symptoms subsided. Overall, they reported a median of one potential symptom (range=0-8) that persisted, with fatigue (40%), muscle aches (22%), alteration of taste (18%), headache (17%), alteration of smell (17%), difficulty in concentration (15%), daytime sleepiness (10%), lightheadedness (7%), and numbness and tingling sensations on the skin (6%), being the symptoms that persisted. Other protracted symptoms reported by subjects were dyspnea (4%), chest pain (3%), and cough (2%). An ANOVA revealed significant differences among the comparison groups (normal, mild and moderate-to-severe PTSD symptoms) regarding the number of protracted symptoms (F (2, 266) = 21.32; p < 0.001). Post hoc Tukey HSD tests indicated that the mean number of protracted symptoms for the moderate-to-severe and mild PTSD groups was significantly higher than no PTSD group.

Separate chi-square analyses (each symptom X PTSD severity) revealed that PTSD severity differed by the presence of each of the above symptoms excluding the alteration of taste and smell, and daytime sleepiness.

### 3.3. Psychiatric status

The psychiatric status of patients and the analyses of these data across comparison groups (normal, mild, and moderate-to-severe PTSD symptoms) are summarized in Table 3. Forty-five patients (15.4%) reported a past diagnosis of a psychiatric disorder. Among these subjects, depression (19 subjects; 6.8%) and anxiety disorders (18 subjects; 6.5%) were the most common diagnoses, and 18 subjects (6.3%) were on psychiatric treatment. A chi-square test (past psychiatric disorder X PTSD severity) showed that PTSD severity differed by the history of a past psychiatric diagnosis (*X*^2^ (2, *N* = 279) = 12.40, p=0.002), and a significantly higher number of subjects reporting moderate-to-severe PTSD symptoms had a past psychiatric diagnosis (p<0.05, post hoc analyses with Bonferroni correction).

**Table 3.** Psychiatric status of patients and the analyses of these data across three comparison groups (normal, mild, and moderate-to-severe PTSD symptoms)

| *Characteristics n (%);*<br>*Mean (SD)* | **Total** | **Normal** | **Mild PTSD**<br>**symptoms** | **Moderate-to-**<br>**severe PTSD**<br>**symptoms** | **X<sup>2</sup>/F** | **df** | **p** |
| --- | --- | --- | --- | --- | --- | --- | --- |
| **Overall** | 284 | 160 (56.3) | 52 (18.3) | 72 (25.4) |  |  |  |
| **Prior psychiatric disorder** | 43 (15.4) | 16 (10.1) | 7 (14.3) | 20 (28.2) | 12.40 | 2 | **0.002** |
| **Impact of Events Scale (IES) score** |  |  |  |  |  |  |  |
| Total | 22.2 (14.8) | 11.2 (5.7) | 27.2 (2.5) | 43.0 (8.5) |  |  |  |
| Intrusion | 7.6 (6.3) | 3.1 (2.4) | 9.8 (2.6) | 15.8 (4.8) |  |  |  |
| Avoidance | 9.4 (5.2) | 6.3 (3.6) | 10.7 (2.8) | 15.4 (4.1) |  |  |  |
| Hyperarousal | 5.1 (4.9) | 1.7 (1.7) | 6.6 (2.3) | 11.7 (3.9) |  |  |  |
| **Hospital Anxiety Depression Scale (HADS)** |  |  |  |  |  |  |  |
| Anxiety score | 6.2 (4.6) | 3.7 [(3.1) | 6.9 (3.7) | 11.1 (4.0) | 101.92 | 2 | **<0.001** |
| Depression score | 6.3 (4.3) | 4.7 (3.6) | 6.5 (3.6) | 9.9 (4.3) | 44.12 | 2 | **<0.001** |
| **Past traumatic events (# of experiences in**<br>**LEC)** | 5.9 (5.2) | 5.2 (4.7) | 7.2 [(5.6) | 6.6 (5.7) | 3.48 | 2 | **0.03** |
| **Pittsburgh Sleep Quality Index (PSQI)** |  |  |  |  |  |  |  |
| Sleep latency score | 1.3 (1.0) | 1.0 (0.9) | 1.4 (0.8) | 1.9 (0.9) | 19.91 | 2 | **<0.001** |
| Sleep duration score | 0.8 (0.9) | 0.6 (0.8) | 0.9 (1.0) | 1.4 (1.0) | 14.89 | 2 | **<0.001** |
| Sleep disturbance score | 1.2 (0.6) | 0.9 (0.4) | 1.5 (0.6) | 1.7 (0.7) | 42.07 | 2 | **<0.001** |
| Subjective sleep quality score | 1.2 (0.7) | 0.9 (0.6) | 1.4 (0.7) | 1.8 (0.7) | 33.14 | 2 | **<0.001** |
| **MINI Suicidality score** | 0.3 (1.2) | 0.1 (0.8) | 0.08 (0.27) | 0.9 (2.1) | 52.17 | 2 | **<0.001** |

Patients reported a median of 4 traumatic life events (range=0-33), and an ANOVA revealed significant differences among the comparison groups (normal, mild and moderate-to-severe PTSD symptoms) regarding the number of traumatic life events (F (2, 240) = 3.48; p < 0.03). Post hoc Tukey HSD tests indicated that the mean number of traumatic events for the mild PTSD groups was significantly higher than no PTSD group.

ANOVAs and post hoc Tukey HSD tests revealed significant differences among comparison groups (normal, mild and moderate-to-severe PTSD symptoms) regarding the PSQI sleep latency score [F (2, 238) = 19.91; p < 0.001; normal < mild PTSD < moderate-to-severe PTSD], PSQI sleep duration score [F (2, 239) = 14.89; p < 0.001; normal, mild PTSD < moderate-to-severe PTSD], PSQI sleep disturbance score [F (2, 238) = 42.07; p < 0.001; normal < mild PTSD, moderate-to-severe PTSD], and MINI suicidality score [F (2, 258) = 9.85; p < 0.001; normal, mild PTSD < moderate-to-severe PTSD].

### 3.4. Outbreak-related variables, media use, stigmatization and social support (Table 4)

One hundred and seventy patients (60.3%) reported that a family member, 51 patients (18.1%) reported that a relative, 40 patients (14.2%) reported that a friend, and 40 patients (14.2%) reported that an acquaintance contracted the COVID-19 infection. Overall, 222 patients (78.7%) reported that someone in the social vicinity contracted the disease. A chi-square analysis revealed that the frequency of the report that someone with the infection existed in the social vicinity was not significantly different between the PTSD severity groups.

**Table 4.**
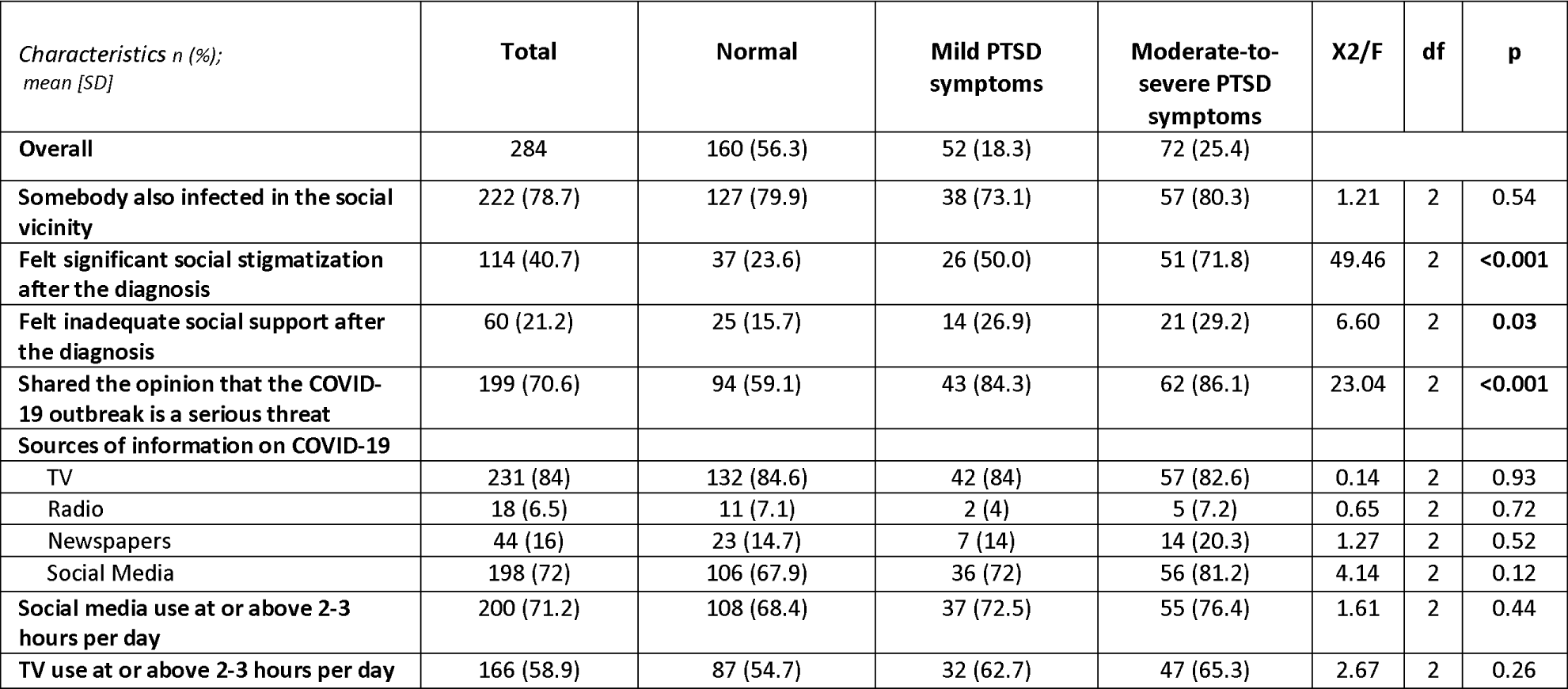
Outbreak-related variables, media use, stigmatization and social support, as well as the analyses of these data across three comparison groups (normal, mild, and moderate-to-severe PTSD symptoms)

The responses of ‘moderately’ or ‘considerably’ to the question on whether the subject felt stigmatized or discriminated against after the diagnosis were scored as significant social stigmatization. Consequently, a chi-square test (significant social stigmatization after the diagnosis X PTSD severity) showed that PTSD severity differed by the report of significant social stigmatization (*X*^2^ (2, *N* = 280) = 49.46, p<0.001), and a significantly higher number of subjects reporting moderate-to-severe PTSD symptoms felt social stigmatization compared to subjects reporting mild PTSD symptoms or no symptoms. Also, a significantly higher number of subjects reporting mild PTSD symptoms felt social stigmatization compared to subjects reporting no symptoms.

The responses of ‘no support’, ‘little support’, and ‘moderate support’ to the question on how much the subject felt social support from family and friends were scored as inadequate social support. Consequently, a chi-square test (inadequate social support after the diagnosis X PTSD severity) showed that PTSD severity differed by the report of inadequate social support (*X*^2^ (2, *N* = 283) = 6.60, p=0.03), and a significantly higher number of subjects reporting moderate-to-severe or mild PTSD symptoms felt inadequate social support compared to subjects reporting no symptoms.

The responses of ‘a very serious threat’ and ‘a serious threat’ to the question on personal view on the seriousness of the COVID-19 outbreak were contrasted against the responses of ‘a small threat” and “not a real threat”. Consequently, a chi-square test (personal view on seriousness of the COVID-19 outbreak X PTSD severity) showed that PTSD severity differed by personal view on seriousness of the outbreak (*X*^2^ (2, *N* = 282) = 23.04, p<0.001), and a significantly higher number of subjects with moderate-to-severe or mild PTSD symptoms considered that the COVID-19 outbreak was a serious threat.

TV (84%) and social media (72%) were reported as the main sources of information on the COVID-19 outbreak in our sample, and daily TV and social media exposure time at 2-3 hours or above was 58.9% and 71%, respectively. The type of media consumed did not differ between the comparison groups. Also, there was not a significant difference between the comparison groups with respect to TV and social media exposure times (F (2, 279) = 1.60; p < 0.20, and F (2, 278) = 1.58; p < 0.20, respectively).

Of the 202 working subjects, 19 subjects (9.4%) reported that they were still on temporary disability leave, and 28 subjects (13.8%) reported that they lost their jobs or were put on temporary leave by the employer during the lockdown. Twenty-seven subjects (13.3%) started working from home or paid infrequent office visits lately, and 128 subjects (63.3%) did not report a significant change in their work routine. A chi-square test showed that PTSD severity in the working subjects did not differ significantly by the state of having lost a job or being on temporary leave by the employer (*X*^2^ (2, *N* = 202) = 0.618, p=0.91). On the other hand, PTSD severity differed by the state of being on temporary disability leave (*X*^2^ (2, *N* = 202) = 6.57, p=0.03), and a significantly higher number of subjects with moderate-to-severe PTSD symptoms (20% of them) was still on temporary disability leave.

### 3.5. Predictors of PTSD symptom severity (Table 5)

The selection of the independent variables for the ordinal logistic regression analysis was based on significance from the univariate analyses (p<0.05), interdependency between the variables, and availability of the data. Among the pre-trauma factors, gender, prior psychiatric disorder, and the number of past traumatic events were entered into the analysis. The subject’s view on the seriousness of his/her acute infection, the number of protracted symptoms, the subject’s view whether the COVID-19 outbreak was a serious threat, social stigmatization after the infection, and the adequacy of social support were the peri-trauma factors that were included. The duration of the acute infectious symptoms was not included, as this variable was not available for all patients (data lacking for 12.7% of the sample). Acute infectious symptom burden (# of symptoms) was also excluded, as this variable had a moderate correlation with the subject’s view on the severity of his/her acute infection (r_s_ = 0.53), another variable which was included in the analysis. Ordinal categories of PTSD symptom severity (no PTSD, mild PTSD symptoms, and moderate-to-severe PTSD symptoms) were the outcome variable. Test of Parallel Lines showed that the ordered probability model met the proportional odds assumption (χ2 = 2.52, P = 0.96).

**Table 5.** Ordinal logistic regression analysis of factors influencing the PTSD symptom severity

| Factors | OR (95% CI) | p |
| --- | --- | --- |
| **Pre-trauma factors** |  |  |
| Female | 2.27 (1.25-4.16) | **0.007** |
| Prior psychiatric disorder | 1.05 (0.46-2.38) | 0.90 |
| Number of past traumatic events | 1.06 (1.00-1.12) | **0.04** |
| **Peri-trauma factors** |  |  |
| Subject's view that his/her acute infection was moderate-to-severe | 1.66 (0.81-3.44) | 0.16 |
| Subject's view that the COVID-19 outbreak is a serious threat | 2.56 (1.21-5.55) | **0.01** |
| Significant social stigmatization | 3.12 (1.69-5.55) | **<0.001** |
| Inadequate social support after the infection | 1.80 (0.90-3.61) | 0.09 |
| Number of protracted symptoms | 1.45 (1.20-1.77) | **<0.001** |

Overall, female gender, increased number of past traumatic events, a personal view that the COVID-19 outbreak was a serious threat (‘very serious’ and ‘serious threat’ versus ‘small’ and “not a real” threat), increased number of protracted symptoms, and significant social stigmatization reported (a rating of ‘moderately’ and ‘considerably’ versus ‘never’ and ‘very little’) were closely associated with increased severity of PTSD symptoms.

### 3.6. Predictors of persistence of symptoms

A multivariable binary logistic regression was performed to determine which independent variables (the epidemiological characteristics, infection-related variables (such as the infection severity), psychiatric status, and outbreak-related variables) were significantly associated with the persistence of the symptoms (one or more protracted symptoms reported by the subject). Selection of the independent variables for the binary logistic regression analysis was based on significance from the univariate analyses (p<0.05) (data not shown), and these variables included gender, prior psychiatric disorder, subject’s view on the seriousness of his/her acute infection, subject’s view whether the COVID-19 outbreak was a serious threat, social stigmatization after the infection, adequacy of the social support, IES-total score, HADS-anxiety score, HADS-depression score, PSQI sleep latency score, PSQI sleep duration score, and PSQI sleep disturbance score.

The logistic regression model was statistically significant, χ2(12) = 48.72, p < 0.001, and it explained 29.3% (Nagelkerke *R*^2^) of the variance in the persistence of the symptoms. Among the variables tested, IES-R total score was the sole independent predictor of the persistence of the symptoms (p<0.001, odds ratio = 1.075 [95% CI, 1.047–1.103]).

## 4. Discussion

We found that a high percentage of patients diagnosed with the COVID-19 infection continued to experience significant psychological distress, after a mean of almost 50 days following the diagnosis. Specifically, around a quarter of the patients in our study reported moderate-to-severe PTSD symptoms, with over 40% of these subjects reporting comorbid depression. Overall, around one-third of the patients in our study reported clinically significant PTSD, anxiety, and/or depression.

Around 40% of the patients reported poor sleep quality in the prior month, a quarter had a sleep duration of 5-6 hours or shorter, and one-fifth had a sleep latency at one hour or longer. Also, one-tenth of the patients had a positive response to at least one of the items of the MINI suicidality scale, which may imply also a heightened risk of suicide, although the risk was ‘low’ in the majority, based on this scale. Finally, one-fifth of the working subjects with moderate-to-severe PTSD symptoms were still on temporary disability leave at the time of the survey.

These findings indicate that a significant proportion of COVID-19 patients may experience psychiatric morbidity in the first few months after the infection. This is in line with the results of the previous SARS and MERS outbreak research, which reported 10% to 35% of psychiatric morbidities in the post-illness stage (Lee et al. 2018). While delirium, insomnia, and symptoms of depression, anxiety, and PTSD have been reported as common features in the acute period of COVID-19 infection, there are few studies investigating the long-term psychiatric status (Rogers et al. 2020). Mazza et. al. reported that more than half of the subjects with prior COVID-19 infection had clinically significant anxiety, depression, PTSD, and/or obsessive-compulsive symptoms, at nearly one-month follow-up after the hospital treatment (Mazza et al. 2020). Similarly, Liu et. al. found that ‘moderate-to-severe’ depression and anxiety were around 10% and 20%, respectively, after about one month following the hospital discharge. In this study, the prevalence of significant PTSD was 12% (Liu et al. 2020). In another study, one-third of the patients with COVID-19 infection reported clinically significant anxiety and/or depression, at a median of 46 days after the virus clearance (Tomasoni et al. 2020).

Limited information exists regarding predictive factors for the mental health problems among COVID-19 patients (Liu et al. 2020). In our study, female gender and perceived stigmatization were significant risk factors associated with the increased severity of PTSD symptoms, consistently with the previous studies (Tolin and Foa 2006; Liu et al. 2020). It is well known that females are not only likely to suffer from depressive and anxiety disorders more frequently, they might also be prone to significant emotional distress and traumatization after major stressors. Perceived stigmatization generally had been related to negative effects on mental health, and had a strong impact on the severity of symptoms after traumas (Mak et al. 2007). In the case of an infectious disease which is highly communicable, and poses a major threat to the public in general, discrimination perceived by the affected individuals might be based on reality, as well as a tendency for self-stigmatization.

Another factor predictive of increased PTSD symptoms were the prior traumatic events reported by the individuals, a finding consistent with the epidemiological studies on PTSD. It is suggested that prior exposure to trauma increases the risk of developing PTSD when a subsequent trauma is experienced (Sareen et al. 2014). We have found that a personal view that the COVID-19 outbreak was a serious threat increased the risk of PTSD symptoms in our sample. Moreover, subjects’ ratings of the severity of their prior COVID-19 infection (i.e., the number of acute symptoms, duration of acute symptoms, and a self-rating of the acute infection severity) were also associated with the PTSD risk, although this association became non-significant after controlling for the effects of other relevant variables. Interestingly, PTSD severity was not related with the objective assessment of the prior infection (i.e. infection severity, lowest oxygen saturation, or supplemental oxygen requirement), and the correlation of these assessments with the individuals’ subjective ratings of his/her acute infection severity were weak to moderate (|r_s_|=0.21-0.52). The above findings indicate that gender and the psychosocial factors such as the prior traumatic experiences, stigmatization, and perceived threat related to the ongoing pandemic, and related to one’s previous COVID-19 infection, rather than the medical factors, may play the major role in the postinfectious psychiatric sequela in COVID-19.

Preliminary findings suggest that a significant proportion of COVID-19 patients continue to experience persistent symptoms, which may result in significant disability (Carfi et al. 2020, Townsend et al. 2020). In our sample, one or more protracted symptoms were reported by 44% of the subjects, with fatigue, muscle aches, alteration of smell/taste, headache, difficulty in concentration, daytime sleepiness, lightheadedness, and numbness and tingling sensations on the skin as the most common symptoms. These are in parallel with the findings of previous studies. Even though the etiology of the persistence of such symptoms is not clear, and this may be heterogeneous, similar symptoms were reported after the onset of other pandemics. In the follow-up of SARS survivors, chronic fatigue persisted 40 months after the infection, and prolonged symptoms and fatigue were present up to 18 months after the MERS infection. (Moldofsky and Patchai 2011; Lee et al. 2019). Interestingly, the strongest predictor of the persistent symptoms, in our sample, was the PTSD symptom severity. Overall, the frequency of moderate-to-severe PTSD in subjects who did not report protracted symptoms was 7.6%, while this was 35.6% for subjects reporting protracted symptoms. This indicates over 4 times higher risk of PTSD in presence of protracted symptoms. A contribution of the posttraumatic psychiatric morbidity on persistence of symptoms is possible, in a substantial proportion of recovered COVID-19 patients.

Clinical similarities between chronic fatigue syndrome and the protracted symptoms after SARS and similar outbreaks have been recognized in the previous decades. (Moldofky and Patcai 2011). Although an ill-defined diagnosis, patients with chronic fatigue syndrome usually present with debilitating fatigue, and other somatic and neuropsychiatric symptoms such as musculoskeletal pain, headaches, sleep disturbances, concentration difficulties, and mood problems. Studies revealed common features between chronic fatigue syndrome and the frequent psychiatric disorders: an overlapping phenomenology, coexistence, shared risk factors, and the neurobiological connections. For instance, a recent large community-based twin registry showed that subjects who reported a history of PTSD were over 8 times more likely to report a history of chronic fatigue syndrome (Dansie et al. 2012). Also, a preceding trauma history, hypocortisolism, and immune alterations are shared by PTSD and chronic fatigue syndrome (Lipschitz and Mo JAMA, 2001). It has been, therefore, proposed that a dysfunctional stress-response system markedly altered by severe and/or chronic stressors (a preceding traumatic life event, or an earlier medical problem such as an infection and its treatment) might be the common pathophysiology for PTSD and chronic fatigue. COVID-19 infection, with its obvious psychosocial dimensions is an area of investigation which may yield important results in the future. Further research aiming to uncover the neurobiological, endocrine, and immune characteristics related to the protracted symptoms and PTSD in patients with COVID-19 is clearly warranted.

There are several limitations to our study. First, we used a convenience sample, and caution must be exercised in generalization of our findings to the broader population of patients with COVID-19. In our sample, the great majority of patients (98.8%) were rated as ‘non-critical’ based on the COVID-19 severity index, hence, further work on the most critical patients is desirable. Second, this was a cross-sectional study which limits our ability to infer causality. Third, this study depended on self-report of patients instead of a structured clinical interview which could provide a better picture of the psychological distress in our patients. Also, a detailed medical examination of the patients with the protracted symptoms could yield additional clues to the cause(s) of persistence of these symptoms, in addition to the psychiatric morbidity. Finally, we did not specifically question the symptom of dyspnea in our protracted symptoms checklist, which may have led to a lower reported frequency of this symptom (4%) in our sample, compared to other studies.

In summary, patients with COVID-19 are prone to substantial psychological distress after the infection. PTSD symptoms and comorbid depression, as well as anxiety, and impaired sleep comprise a substantial part of the distress described by these individuals. Various personal (i.e., gender and prior trauma history) and psychosocial factors (i.e., perceived stigmatization and a personal view on seriousness of the threat posed by the COVID-19 pandemic) are likely to mediate the mental health effects in the context of COVID-19. The protracted symptoms are also frequent in this period, and these symptoms are related to the posttraumatic psychiatric morbidity.

## Data Availability

Data is available to researchers in case of request.

## Acknowledgments

We thank our patients for their readiness to engage in this study.

